# Community and health system factors influencing snake envenomation management practices in three districts of Ghana: a qualitative inquiry from health stakeholders and snakebite victims

**DOI:** 10.1101/2023.02.16.23286015

**Authors:** Mabel Worlasi Dzenu, Afaya Agani, Martin Ayanore

## Abstract

**Introduction:** Snakebite is a neglected public health issue affecting individuals of all ages in many tropical countries. Venom from snakebite is a potentially life-threatening disease associated with severe morbidities and mortalities. However, literature on snakebite envenoms management practices remains understudied. This study sought to explore community and health system factors influencing snake envenomation management practices in three districts in the Oti region of Ghana. Also, we assessed the current adherence to snakebite treatment protocols to the national and World Health Organization (WHO) recommended protocols.

**Methods:** The study adopted an exploratory descriptive qualitative study design. Purposive sampling technique was used to recruit participants for the study. The participants included snakebite victims, traditional healers, and health professionals. In-depth interviews and focused group discussions were used to collect data from the participants. All the interviews were audio recorded and transcribed verbatim and exported to Atlas Ti for data management and analyses. The transcribed data were analyzed using the content analysis method to develop the categories for the study findings

**Findings:** Twenty-six (26) participants were recruited including 16 males and 10 females. The study found three main factors that influenced snake envenomation management practices. These factors included health system factors, community-level factors, and adherence to national and WHO-recommended protocols. The health system factors included the unavailability of anti-snake venom, high-cost ASV, and inadequate or no supply of ASV from Ghana Health Service. The community factors included delay in seeking treatment, transportation, and accessibility challenges, and non-community acceptance facility treatment of snakebites. Almost all the health care professional who provide care for snakebite victims adhered to the WHO recommendations.

**Conclusion:** The evidence adduced from this study could be used by public health practitioners to empower the community by increasing awareness of the prevention of snakebites, first aid, and appropriate treatment-seeking behavior. Culturally appropriate interventions that seek to increase awareness to improve the acceptability of hospital management and sensitize the community for early referral of snakebite victims to the nearest health facility having anti-venom treatment.

**Authors summary:** The negative responses by the community’s acceptance of the kind of treatment given in the health facilities and the unavailability of Anti-snake Venom (ASV) in the study districts’ health facilities inhibit the management of snakebites.

## Introduction

Snakebite is a neglected public health issue affecting individuals of all ages in many tropical countries [1, 2]. Venom from snakebite is a potentially life-threatening disease associated with severe morbidities and mortalities [3, 4]. Among countries in the tropical and subtropical areas, snakebite has been regarded as a neglected public health issue necessitating rapid attention [1]. Though the exact number of snake bites is unknown, an estimated 5.4 million snake bites occur yearly, resulting in 1.8 to 2.7 million cases of envenoming (poisoning from snake bites) [2]. Out of this, the fatality rate ranged between 81400 and 137880, and around three times as many amputations and other permanent disabilities have been recorded each year [2].

Snake envenomation is one of the most significant human-versus-wildlife conflicts. Data on the economic burden of snake bites globally is scarce [2]. The cost of clinical care is high for snake bite victims and their families [5] in low- and middle-income countries. Undoubtedly, vulnerable populations experience catastrophic health expenditures and poor quality of life (QoL), particularly in healthcare systems that do not prioritize and provide timely support and care to ameliorate the cost of antivenom treatments. The economic and societal cost of snakebites is substantial in underdeveloped countries. Over 2.8 million people are bitten by snakes each year in India, with approximately 46,900 people dying as a result. Around 589,919 snake bites occur in Bangladesh each year, with 6041 resulting in death [6].

Up to a million individuals are claimed to be bitten each year, with estimates of 7000-20,000 deaths per year [7]. West Africa alone has 3557-5450 fatalities. According to a WHO report, 6687 snakebites were treated in one hospital in Nigeria in just three years, while in Burkina Faso, 114 126 snakebites were reported nationally over five years from 2010 to 2014 [6].

One major challenge that has confronted the effective case management and clinical care for victims of snakebites is the reliance on ineffective traditional medicine as an alternative treatment by many people, particularly in developing and low-resource settings. Empirical studies across sub-Saharan Africa (SSA) showed a high proportion of snakebite victims who resort to the use of traditional medicine for treatments. In Mali, up to 49.7% of snakebite victims seek treatment from traditional sources. In Nepal, 56% of victims reported traditional medicine as primary health-seeking behavior. In Kenya, the figure is at least 68% [6]. Many population groups in SSA resort to the use of traditional medicine, which ends up being ineffective. The crisis in access to safe, effective anti-venom treatment in some country contexts. Poor access to health services, limited infrastructure, poverty, and cultural barriers contributed immensely to morbidity and mortality [8]. In Ghana, an average of 9,600 snakebites were recorded each year from 2015 to 2019 [9]. A community-based study found a high snakebite prevalence of 6% and a case-fatality ratio of 3% [10]. Case fatality rates in one health facility fell from 11.1% to 1.3% 33 months after implementing a review and treatment system [11, 12]. The WHO has called for country studies to address knowledge gaps to support country health systems in effectively addressing this neglected health problem of snakebites [1]. Findings from this study identified community and health systems factors that will help with policies for treatment systems to effectively support snakebite envenoming and improved long-term support and health outcomes of treatment. This study, therefore, sought to address this knowledge gap.

## Methods

### Study site description

The study was conducted in three snakebite endemic districts; Nkwanta North, Krachi Nchumuru, and Krachi West in the Oti Region, The Oti Region is one of the six newly created regions formed out of the previous ten. It was created by the Constitutional Instrument (C.I) 112 after an undisputed majority affirmed its creation in a referendum. The region is predominantly made up of the Guan ethnic group with the second majority being Ewes.

The region has a much arid climate as compared to the southern areas of Ghana due to its proximity to the north. The average annual rainfall varies from 750 to 1050 mm. The main occupations include farming (yam, maize, and cash crops), trading, and fishing while others are employed in state-owned and private institutions.

#### Design

The study employed an exploratory descriptive qualitative approach. The design was appropriate as it helped to unearth and develop a deeper understanding of the community and health system factors influencing snakebite treatment and management practices. Additionally, this study design allowed us to be able to answer questions about why and how snakebite victims are managed in the Oti region of Ghana.

#### Participants recruitment and sampling

Participants for the study were snakebite victims, traditional healers, Disease Control Officers, District Director of Health Services, Regional Director of Health Services, drug shop owners or pharmacists, and health staff (nurses). Purposive sampling technique was used to recruit the participants for the study. These participants were included in the study due to their prior in-depth information on the phenomenon under study. All eligible participants who consented were included in the study. For health workers, only those living or working in the three districts; Krachi West, Krachi Ntsumuru, and Nkwanta North were included. Again, snakebite victims who did not consent were excluded from the study. The study also excluded traditional healers who treat other conditions besides snakebites in the three districts. All participants who were seriously sick or showed severe signs of COVID-19 at the time of the data collection were excluded from the study. The sample for the study was determined using the saturation method [13]. With this, the interviews and transcription were done concurrently.

Prior to the data collection, preliminary visits were made to the study sites to first recruit the participants for inclusion. The participants obtained during the visit were provided with information sheet and informed consent form. Participants who willingly agreed to participate in the study signed the informed consent form and the date for the interviews was discussed with time. The participants were given permission to select the preferred day within the data collection period for the interviews to be conducted.

#### Data collection and procedures

Three research assistants were trained prior to the data collection. The training spanned three days. It covered the aim of the study, consenting process, and the procedures for data collection. A semi-structured interview guide was used to collect qualitative data from the participants. Existing literature in the setting influenced the construction of the interview guide [14]. We developed three interview guides (one for victims, one for traditional healers and drug shop owners or pharmacists, and one for regional and district directors and disease control officers of health), and one FGD guide which were all pre-tested and adapted after two pilots.

In-depth interviews and FGDs in each study district were conducted, either in a quiet secluded room (in-depth interview) or a private meeting room (FGD). The in-depth interviews were between 10-30 min in duration and FGDs circa 60 min or more. However, all the interviews were conducted in English and audio-recorded for later transcription while the data collection commenced in January and ended in June. When the intended theoretical sample size was reached, themes appeared, and full data saturation was achieved, data gathering was judged complete.

#### Data Analysis

We transcribed and analyzed all the in-depth interviews and FGDs. Content analysis was used for this study since it aids in generating insights from textual data [15]. To do this, all transcripts were exported to Atlas. ti for in-vivo coding [16]. Two researchers independently coded three transcripts before discussing and agreeing on a coding scheme [17]. One researcher coded all remaining transcripts, with further iterative adaptations to the coding scheme discussed and agreed upon by two researchers. We explored relationships between codes, similarities, and conflicts within and between individual participants and checked for patterns across the whole dataset and by participant and facility characteristics to identify and construct themes. We carried out reputational searches, checking for any disagreement with proposed patterns and themes. The qualitative analysis was done to answer community and health system factors that facilitate or inhibit treatment outcomes for snakebite victims and snakebite treatment protocols in the study. Data with the same codes were gathered and organized into themes to obtain a clear overview of the different responses [18]. The results were presented using a narrative approach (extracts and quotes from the themes and sub-themes generated were used to support the results of the study).

#### Rigour

With the due importance accorded rigour or trustworthiness in qualitative research, therefore, we made an effort to guarantee confirmability and transferability. Due to the thorough explanation of the study’s methodology, the results could be transferred and verified. We used three participants for member checking to ensure the credibility of the findings. This was done a week after transcription was completed so that participants could verify the transcripts accurately matched the interview material. The interviews were read again to ensure the legitimacy of the results, and the comments of colleagues were frequently used. External oversight was also used to boost its legitimacy. A portion of the data was sent to a researcher unrelated to the study as a foreign observer to see if he or she had a comparable understanding of the data. Strategies such as rewriting, member check, examining texts and using the participants’ own words, monitoring translations, capturing audio, conducting the interview, coding, themes and sub-themes, and the use of direct quotes to support the conclusions were utilized to promote rigour [19]. To ensure data transferability, the setting of the investigation and the description of the phenomena observed were specified for comparison. To ensure reliability, the study procedure was clearly defined so that other researchers may undertake the same investigation.

#### Ethical approval

Ethical approval for the study was obtained from the Ghana Health Service Ethics Review Committee (GHS-ERC: 019/03/21). Institutional permission was sought from the Volta Regional Health Directorate, and the heads of the various facilities included in the study. Written informed consent was sought from every participant before inclusion into the study. All the participants in the study were given an information sheet, which contained information on the study’s aim, methods, potential risks and benefits, compensations, confidentiality, and anonymity as well as the contact information of the principal investigator. The participants who agreed to participate signed the informed consent. All transcripts and audio recordings were anonymized by being given pseudonyms in order to eliminate any personal information that may be used to identify the study participants. To prevent unauthorized individuals from accessing the material, we additionally encrypted a passcode and locked the audio files and transcripts.

### Participants and public involvement

The study participants and the public were not involved in the design of the study. Snakebite victims, health professionals, and traditional healers participated in the study as participants. Findings from the study are intended to be shared with members of the public, healthcare professionals, and relevant stakeholders via scientific publications, lay reports, social media, and conferences both within Ghana and beyond.

## Findings

### Participants demographics characteristics

This study involved in-depth individual interviews that included thirteen (18) participants; two (4) females and eleven (14) males while the FGD included twelve (12) participants; five (6) males and eight (8) females. The total individual participants sum up to; twenty (31) participants. However, (8) participants; District Disease Control Officer and Health Director and 6 Staff Nurses were recruited from Krachi Ntsumuru District, (7) participants; District Disease Control Officer and Health Director and Snakebite victims from Nkwanta North District, (9) participants; District Disease Control Officer and Health Director and Emergency Unit staffs (Nurses) and the Regional Health Director.

**Table 1.1.** Demographics of study participants.

| **In-Depth Interviews** | **Participants N= (31)** | **SEX** |
| --- | --- | --- |
| Snakebite victims | 5 | 2 Females<br>3 Males |
| Regional director and District Health directors | 4 | 0 Females<br>4 Males |
| District Disease Control officers | 3 | 0 Females<br>3 Males |
| Drug shop owner (pharmacist) | 1 | 0 Female<br>1 Male |
| Snakebite Victims | 5 | 2 females<br>3 males |
| **FOCUS GROUP DISCUSSIONS (FGD)** |  |  |
| Health Staff (Nurses) | 6 | 4 Females<br>2 Males |
| Emergency Unit staff (Nurses) | 7 | 4 Females<br>3 Males |

#### Health system factors influencing the treatment of snake bites

This study identified some health system factors that influence the treatment of snake bites within the district health facilities. One of the key factors identified was the unavailability of Anti-Snake Venom (ASV) for the treatment of snake bites in health facilities within the districts. Some participants mentioned that there is an inadequate supply of ASV to the districts which makes the management of snake bites challenging. While some participants mentioned that the government has completely stopped the supply of ASV which makes the procurement of these ASV very challenging due to the high cost. A participant narrated:

> “*…government do supply all these things, but for some years now it appears as if the government has STOPPED and the cost of procuring this anti-snake is very high. So, this issue of government supplying or supporting the anti-snake, fight, or treatment has ceased and this is serious…There is no support. There is no donor support or any support that is in place to support anti-snake or the provision”*.
>
> *- (In an interview with one of the district officers)*

Even though most of the participants stated the unavailability of the ASV, some felt the supply of the ASV has ceased from GHS while others said they received only a few. Some participants reported that the supply of ASVs from the national through the regional level to the district level comes in small quantities, where snake bite victims are forced to buy them elsewhere for treatment. A participant narrated:

> “…*unavailability of treatment is also another challenge because I know it takes some time for them to get the ASV and I think they have a quarter that comes from National to Regional and to Districts but in a very minute quantity, so people are forced to buy it from the open market and keep it somewhere and be selling out to the public…”*
>
> *- (in an in-depth interview with one District officer)*.

Another participant said during the IDI started that it is not a problem when you are supported with the ASV during the snakebite treatment because they are unavailable, but you have to buy at a subsidized price. He stated that they have not received any donation or support as a district. The legislator (Member of Parliament) provided ASV that they could buy at a low cost.

> “*The main challenge is the unavailability of ASV and the costs of it, is not a problem if you are supported. The ability of the people to pay is one of the challenges we face here*… *I can say that the MP supported with ASV at a subsidized price” - (one of the directors during an IDI)*.

#### Community-level factors that affect the treatment of snakebites

This study observed some community-level factors that affected the treatment of snakebites. Some participants claim that the community’s delay in seeking healthcare hinders the outcome, while others claim that transportation and accessibility challenges to the institution for treatment, as well as financial issues, hamper the outcome. Some participants indicated that some communities do not even accept the treatment provided by the hospital due to financial restrictions; they further stated that some community members prefer to seek treatment from traditional healers in terms of snakebite rather than seeking care from the hospital. A participant stated:

> *“They wouldn’t accept it, because it involves money and the people don’t have the money and their level of education is as low as you can think of. Yes, because it’s unacceptable for somebody to carry 12 million and go and take only anti-snake. When they know very well that at the local place when you go, you may not even pay a pin. So why should I go to a government hospital, so they don’t accept it…”*
>
> *- (in an in-depth interview with one District officer)*

Nonetheless, some of the participants also believed that they know they are supposed to go to the hospital for treatment, they prefer the traditional healers and they do not accept the treatment given in the hospital. A victim stated:

> “*…I don’t think they don’t accept it considering the amount of you will spend and the days as compared to our people at home. Even though we are to go to the hospital, we prefer to go to our people”*
>
> *- (in an in-depth interview with one snakebite victim)*

### Adherence to current national and WHO-recommended protocols for snakebite treatment

It was revealed that almost all participants in the facilities that treat snake bite follows the WHO-recommended protocol guidelines for the treatment of snakebite. Some (7) of the participants reported having and following the recommended protocols for the treatment and management of snake bites while some participants (10) reported that they do not have the protocol guidelines pasted but they have it at the facility level where treatment is done and they follow it strictly. However, some (3) say they do not have the recommended protocols but will look forward to providing that in the future. One of the participants stated that;

> *“Yes, we don’t have the protocol pasted here because we don’t do treatment. But at the facility level where and this one is managed by either a medical doctor or a trained staff nurse… So, at the facility level, where they manage snakebite, the protocol is there and they adhere to it…”*
>
> *- (in an in-depth interview with one District officer)*

The above statement was supported by 3 others during a FGD in the health facility that, they have the WHO protocol guidelines at the facility that they follow to do their treatment and management all the time.

> *“Yes, we were having but when they were doing renovation, they took it off but we use the protocol recommended by WHO for the treatment”*
>
> *-(R1, R2, F5 (FDG participants)*

Nonetheless, while others had the protocol and are adhering to it, some of the participants stated, they do not have the protocol but will in the future;

> *“No, for now, we don’t have any protocol on snakebite in the facilities, and maybe going forward that’s what we should do, we should look at getting a protocol at the facility level for them to follow when there is snakebite for them to be able to manage it”*
>
> *- (one of the directors during an IDI)*.

This was supported by almost all the participants, which means that they know the guideline protocols to follow during treatment.

## Discussion

This study explored the community and health system factors influencing snake envenomation management practices in three districts within the Oti region of Ghana. The study found that health system factors and community-level factors influenced the management of snakebites within the Oti region.

Sub-Saharan Africa has the second largest burden of snakebites after Asia, but there is only one producer of ASV based in South Africa [20, 4, 21]. Several producers of varying amounts of ASVs for use in SSA are based in Europe, Asia, and Latin America which affects the supply chain. While ASVs are relatively available in other endemic areas in Latin America and Asia, SSA has faced major challenges with a reliable supply of effective products of ASVs [22]. This study found that the unavailability of ASVs in health facilities affected the management of snakebites. Our finding is consistent with a recent study conducted in three sub-Saharan African countries (Kenya, Uganda, and Zambia) where there was a lack of ASVs in most health facilities [23]. Also, another study in Cameroon found that there was unavailability of antivenom in health facilities [24]. The plausible reason for this finding could be that in 2016, there was a cessation of the production of excellent antivenom used in the region. This finally served to galvanize the global health community into action toward the control of snakebite envenomation, especially in developing countries [22]. We could also explain that within developing countries resources are generally scarce to sustain antivenom availability in the long term, and rural facilities are usually not provided with sufficient antivenom supplies [25]. Also, possibly the current deployment approach of providing ASV in urban tertiary facilities fails to reach remote health facilities. We suggest that there is a need to strengthen the logistics of procurement, distribution, and monitoring, as well as forecasting and rational utilization of ASVs.

This study found that snakebite victims delayed in reporting to health facilities for treatment. The delay in reporting to health facilities was particularly due to prehospital treatment where most participants preferred traditional treatment. This finding is consistent with a study conducted in India where snakebite victims were reluctant to visit government hospitals but preferred traditional or alternative treatment [26]. Evidence shows that traditional remedies have not been proven to be effective in treating snakebites, so therefore it is recommended to avoid traditional first-aid treatment and alternative medical/herbal therapy [27]. A study conducted in Cameroon revealed that snakebite victims who received traditional treatment was strongly associated with envenoming severity and complications [24]. We, therefore, recommend public health programs that support community awareness and improve coordination between health facilities and traditional healers to allow quick referral of several cases to the hospitals.

Nonetheless, the non-acceptability of the community on the kind of treatment given and challenges or limitations faced during treatment and management of snakebite. The non-acceptability was found because of financial constraints and the high cost of treatment. Lowering the price of antivenom treatment shows to be cost-effective and also increases the acceptability of the treatment now given [28]. The healthcare and the required treatment cost of treatment of snakebites determine the type and place seek healthcare. The cost of antivenom (as well as the cost of supportive care) influence heavily the total cost of hospitalization, which in turn influences the cost-effectiveness of snakebite interventions [29]. The health economic impact of snakebite treatment estimates the cost of managing snakebite envenoming at the local hospital level since victims are responsible for the cost [30, 28]. Antivenoms are frequently inaccessible to both patients and the suffering healthcare system due to their high cost [31].

The adherence of snakebite treatment protocols in the study districts were found to be low in the current treatment; some facilities also reported not to have the recommended guideline protocols resulting to low adherence to the treatment protocol for snakebite management and treatment while some have the guidelines protocol and follow it strictly. [32] reported that the unavailability of any recommended snakebite guidelines protocols for treatment of snakebite at the health facilities contribute to treatment outcome. Furthermore, [2] the WHO guidelines recommend that all clinicians should urgently decide to administer ASV following a snakebite. Low adherence to national standards for patient treatment and a paucity of antivenom suitable for African dangerous snakes are both problems [33]. Similar results were found by [34] that, healthcare providers follow the recommended WHO protocol for the treatment of snakebite.

Additionally, per the recommendations of WHO, the most effective treatment for snakebite is the administration of monospecific anti-snake venom with the recommended protocol guideline for treatment [35, 1]. However, this therapy is not always available to snakebite victims because of some facilities do not have the WHO recommended protocol guidelines. To a large extent the manifestation of snakebite depends upon the species of snake, and therefore identification of the type of snake is important [30]. At times the bite mark might not be visible. The killed snake brought as evidence helps in identification of snake, in which case species-specific monovalent antivenom can be administered. The clinical manifestations of the patient may not match with the species of snake brought as evidence. It is therefore advantageous to know the appearance of the snake so as to recognize the species [30, 4].

### Implication for health system strengthening and practice

Since the current deployment strategies of antivenom mostly rely on providing small stocks to hospitals especially in rural endemic areas, it is imperative that hospital staff to ensure periodic request replenishment from the national headquarters. We suggest that to increase accessibility the ‘hub-and-spoke distribution method is imperative, wherein facilities in rural areas serve as satellites or spokes and are linked tertiary hospitals in urban areas for referrals and for more readily available antivenom supplies [22]. Also new approaches for delivering antivenom are increasingly being considered for remote and inaccessible areas. For example, in well developed countries helicopters have been used to deliver lifesaving ASV and interventions. Due to the financial implications of the delivery method in low-income countries, we suggest that to deliver life-saving ASVs in remote areas [36].

We recommend that further research is required to determine the most appropriate, simple, practical, affordable, and scalable approaches for delivery and deployment of snakebite care ASVs for rural areas.

#### Limitations and suggestions

The study was heavily dependent on the skills of the researchers to get in-depth information from the participants and the results can not be generalized and statistically representative. There may be recall bias from the respondents and social desirability bias on the requested data. However, participants were made to review their results and multiple coding was done by all the researchers. We suggest that during qualitative studies; the objectivity of the study should be maintained and bias should also be avoided during selection and data analysis as much as possible.

## Conclusion

This study adduces evidence on the health system and community level factors that influence the management of snake bite victims. The findings could be used by public health personnel to empower the community by increasing awareness on the prevention of snakebite, first aid, and appropriate treatment-seeking behavior. Culturally appropriate interventions that seeks to increase awareness to improve acceptability of hospital management and sensitizing the community for early referral of snakebite victims to the nearest health facility having anti-venom treatment.

## Data Availability

The data will be made available upon the request. this is because it is a qualitative reseacrh and the data are in form of words in several documents (transcripts) that were analysed and represented in the results.

